# Family screening for hypertrophic cardiomyopathy: initial cardiologic assessment, and long-term follow-up of genotype-positive phenotype-negative individuals

**DOI:** 10.1101/2024.11.01.24316619

**Authors:** Stephan A.C. Schoonvelde, Georgios M. Alexandridis, Laura B. Price, Arend F.L. Schinkel, Alexander Hirsch, Peter-Paul Zwetsloot, Janneke A.E. Kammeraad, Marjon A. van Slegtenhorst, Judith M.A. Verhagen, Rudolf A. de Boer, Michelle Michels

**Affiliations:** Department of Cardiology, Cardiovascular Institute, Thorax Center, Department of Cardiology, Erasmus Medical Center, Rotterdam, the Netherlands; Department of Radiology and Nuclear Medicine, Erasmus Medical Center, Rotterdam, the Netherlands; Netherlands Heart Institute, Utrecht, The Netherlands; Department of Pediatric Cardiology, Erasmus Medical Center – Sophia Children’s hospital, Cardiovascular Institute, Rotterdam, the Netherlands; Department of Clinical Genetics, Erasmus Medical Center, Rotterdam, The Netherlands

**Author notes:** Corresponding author: Stephan A.C. Schoonvelde, MD Department of Cardiology, Cardiovascular Institute, Thorax Center, Erasmus MC Dr. Molewaterplein 40, 3015 GD Rotterdam, The Netherlands.

**Keywords:** hypertrophic cardiomyopathy, pre-symptomatic testing, genotype, phenotype, echocardiography

## Abstract

**Aims:** (i) Investigate the prevalence of hypertrophic cardiomyopathy (HCM) in individuals with pathogenic/likely pathogenic (P/LP) gene variants detected through family cascade testing in relatives, and (ii) evaluate phenotypic progression in genotype-positive phenotype-negative (G+/P-) individuals during follow-up.

**Results:** From 2000-2023, 273 individuals underwent cardiologic evaluation following P/LP variant detection through family screening. Upon initial evaluation, HCM was diagnosed in 128 (47%) individuals. Comparing with 145 G+/P- individuals, HCM patients were older (48 vs 38 years, *p*<0.001) and more likely male (57% vs 34%, *p*<0.001). During follow-up (median 11 years), 14 (11%) of the HCM patients died (two from sudden cardiac death), four (3%) underwent myectomy, 15 (12%) developed atrial fibrillation and 17 (13%) required implantable cardioverter-defibrillator implantation (15 primary prevention, 88%). HCM-related adverse outcomes correlated with younger diagnosis age.

During follow-up (median 8 years), out of the 118 G+/P- subjects, seven (6%) individuals (71% female, diagnosed age 39-77, after median follow-up 6 years) developed HCM (mean maximal wall thickness increasing from 10.2 mm to 13.3 mm). In this G+/P- cohort, significant echocardiographic changes from baseline to last visit were negligible. Over half (56%) had <1 mm change of maximal wall thickness. No adverse cardiac outcomes occurred.

**Conclusion:** The initial evaluation was high-yield, with HCM being diagnosed in 47% of G+ individuals, more frequently in older males. Over a median 8-year follow-up, 6% of G+/P- individuals developed mild HCM, with no adverse cardiac outcomes. These data support initial screening in all first degree relatives, but (very) low-frequency cardiologic evaluations for G+/P- individuals thereafter.

## Introduction

Hypertrophic cardiomyopathy (HCM) is the most prevalent inherited cardiac disease, with an estimated prevalence of 0.2-0.5%. It is characterized by left ventricular (LV) hypertrophy (LVH) in the absence or without excess of abnormal loading conditions. [1-3] Approximately half of HCM patients carry a pathogenic or likely pathogenic (P/LP) sarcomere gene variant. Identifying these P/LP variants enables the detection of relatives at risk of HCM. Current guidelines recommend cascade genetic testing of first-degree relatives in families where a P/LP gene variant is identified. [4-6] Pre-symptomatic testing (PST) aims to identify these at-risk relatives through genetic testing, followed by cardiologic evaluation. [7] Current guidelines recommend cardiologic screening, including electrocardiography (ECG) and cardiac imaging, for all first-degree relatives with an identified P/LP variant to detect the presence of an HCM phenotype. [7, 8] Given that asymptomatic HCM patients are also at risk of disease progression and sudden cardiac death, such screening is important for the early detection of HCM. [9-11]

To date, the specific factors influencing the disease penetrance of HCM in P/LP gene variants remain incompletely understood. [12-14] Disease penetrance is age-dependent, warranting repeat evaluations. HCM is rare in children and more prevalent in males, who are diagnosed at a younger age compared to women. [6, 10, 15, 16] Not every individual with a P/LP gene variant will develop HCM due to an incomplete penetrance. Predicting whether asymptomatic individuals will develop a phenotype is challenging. Cardiologic screening in relatives who carry a P/LP variant but do not exhibit the HCM phenotype, referred to as genotype-positive, phenotype-negative (G+/P-) individuals, is currently recommended every 1–3 years before age 60 and every 3–5 years thereafter according to European guidelines, and every 3–5 years in all adults per American guidelines. [7, 8] Previous studies have shown minimal and infrequent progression to overt HCM in G+/P- individuals within 4-7 years of follow-up. [14, 17-19].

The aims of this study were to (i) assess the prevalence of HCM upon the first cardiologic evaluation in subjects with a P/LP gene variant and (ii) to analyze the evolution of echocardiographic parameters and the development of HCM during long-term follow-up of G+/P- individuals.

## Methodology

A retrospective analysis was performed which included individuals from a single expertise HCM referral center (Erasmus MC, Rotterdam, the Netherlands). The study was conducted according to the principles of the Declaration of Helsinki. All individuals gave written informed consent for inclusion in the local registry for which Institutional Review Board approval was obtained. We included all G+ individuals with HCM-associated P/LP genes identified during family screening referred for cardiologic examination. Gene variants were interpreted using the 2015 American College of Medical Genetics and Genomics/Association for Molecular Pathology guidelines. [20] HCM was diagnosed according to the guidelines, based on a maximal wall thickness (MWT) ≥13 mm on echocardiography in G+ individuals. LV outflow tract (LVOT) obstruction was defined by the presence of a maximal LVOT gradient of ≥30 mmHg. Baseline differences between G+ individuals diagnosed with HCM, and G+/P- were described. A history of hypertension was defined as a prior diagnosis of hypertension that was known and managed with medical therapy at the time of cardiologic screening. HCM-related outcomes were reported, which included the development of atrial fibrillation, implantable cardioverter-defibrillator (ICD) implantation, and appropriate ICD therapy; septal reduction therapy, and (sudden) cardiac death. A composite adverse cardiac event variable was defined by the occurrence of any of these events.

The second analysis focused on the long-term longitudinal follow-up of G+/P- individuals. For this analysis, individuals with at least two ECGs and transthoracic echocardiograms performed more than one year apart were included. All baseline, and most recent follow-up ECGs and echocardiograms were reassessed. Indexed left atrial volumes were retrospectively calculated from the images using the biplane Simpson’s method. [21] Diastolic function was evaluated using the 2016 American Society of Echocardiography/European Association of Cardiovascular Imaging diastolic function assessment guidelines. [22] ECGs were visually assessed, and significant change was defined as any (i) development of a non-sinus rhythm, (ii) abnormal prolongation of conduction times, (iii) significant alterations in QRS morphology (ie, fragmentation, abnormal Q wave or bundle branch block development), (iv) deviation (≥1 mm) of the ST-segment in more than one lead, (v) alterations in T-wave morphology in more than one lead and/or (vi) the presence of LVH voltage criteria (based on any of the Romhilt– Estes, Sokolow-Lyon and Cornell criteria).

Statistical analysis was conducted using IBM SPSS Statistics version 28 (SPSS, IBM, Armonk, New York). Unpaired data was analyzed using the independent samples t-test for normally distributed continuous data, the Mann-Whitney U test for non-normally distributed continuous data, and the chi-square test for categorical data. Paired data was analyzed using the paired sample t-test for normally distributed data and the Wilcoxon signed-rank test for non-normally distributed data. Normality of data was assessed using the Kolmogorov-Smirnov test. Logistic regression was utilized to correct for baseline differences with respect to age. Tests were two-sided and statistical significance was assumed for *p*<0.05.

## Results

### Diagnostic yield of cardiologic examination

Between November 2000 and August 2023, 273 individuals (45% male, mean age 44 years) were referred for cardiologic examination after PST identified a P/LP HCM-related gene variant. An overview of the entire cohort and the main findings are illustrated in Fig. 1. The most common P/LP variants were identified in *MYBPC3* (214; 78%) and *MYH7* (23; 8%). At initial cardiac evaluation, 128 individuals (47%) were diagnosed with HCM (mean MWT 14.7, range 13-24 mm). Individuals diagnosed with HCM (as described in Table 1) were predominantly male (57% vs 34%, *p*<0.001), older (48 vs 38 years, *p*<0.001), and more likely to have a history of hypertension (23% vs 9%, *p*=0.002) than G+/P- subjects. As represented in Fig. 2, the prevalence of HCM increased with ascending age. After correcting for age at diagnosis, a history of hypertension did not remain a significant contributor (*p*=0.311) for having a HCM phenotype (odds ratio [OR] 1.04 per year, 95% confidence interval (CI) 1.02-1.05, *p*<0.001). Body mass index (BMI) did not significantly differ between patients diagnosed with HCM and G+/P- individuals (26.3 vs 25.1 kg/m^2^, p=0.069). Most HCM patients had a variant in *MYBPC3* (106; 83%), followed by MYH7 variants (12; 9%). In the remaining patients (10; 8%), other HCM-associated gene variants were found. One HCM patient had a P/LP variant in both *MYBPC3* and *MYL2*. The youngest G+ individual diagnosed with HCM was 10 years old. A total of three G+ individuals were diagnosed with HCM before the age of 16 years, all with *MYBPC3* variants. A further 35 (27%) patients were diagnosed after the age of 60, with the oldest being 82 years. There was no significant difference in the age of diagnosis between the different genes in this cohort.

**Figure 1.**
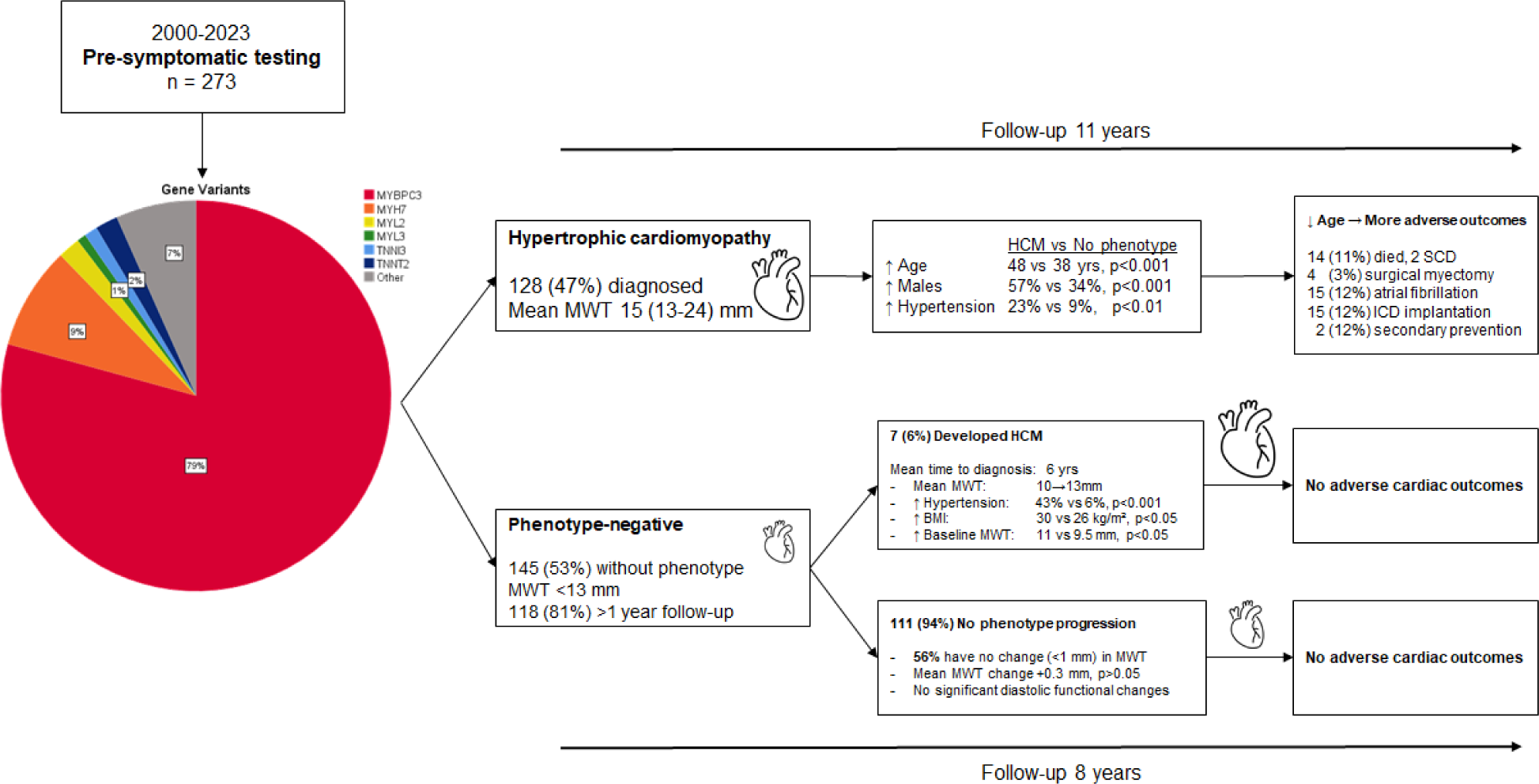
Schematic overview illustrating the genetic yield from pre-symptomatic testing, the diagnostic yield following cardiologic screening, and the key differences between individuals diagnosed with hypertrophic cardiomyopathy (HCM) and those without a phenotype at initial screening. The diagram also highlights findings in initially phenotype-negative individuals who later developed HCM, the progression of the phenotype in those who remained phenotype-negative, and the reported adverse cardiac outcomes across these three distinct groups. <u>Abbreviations</u>: BMI: body mass index; ICD: implantable cardioverter-defibrillator; MWT: maximal wall thickness; SCD: sudden cardiac death.

**Figure 2.**
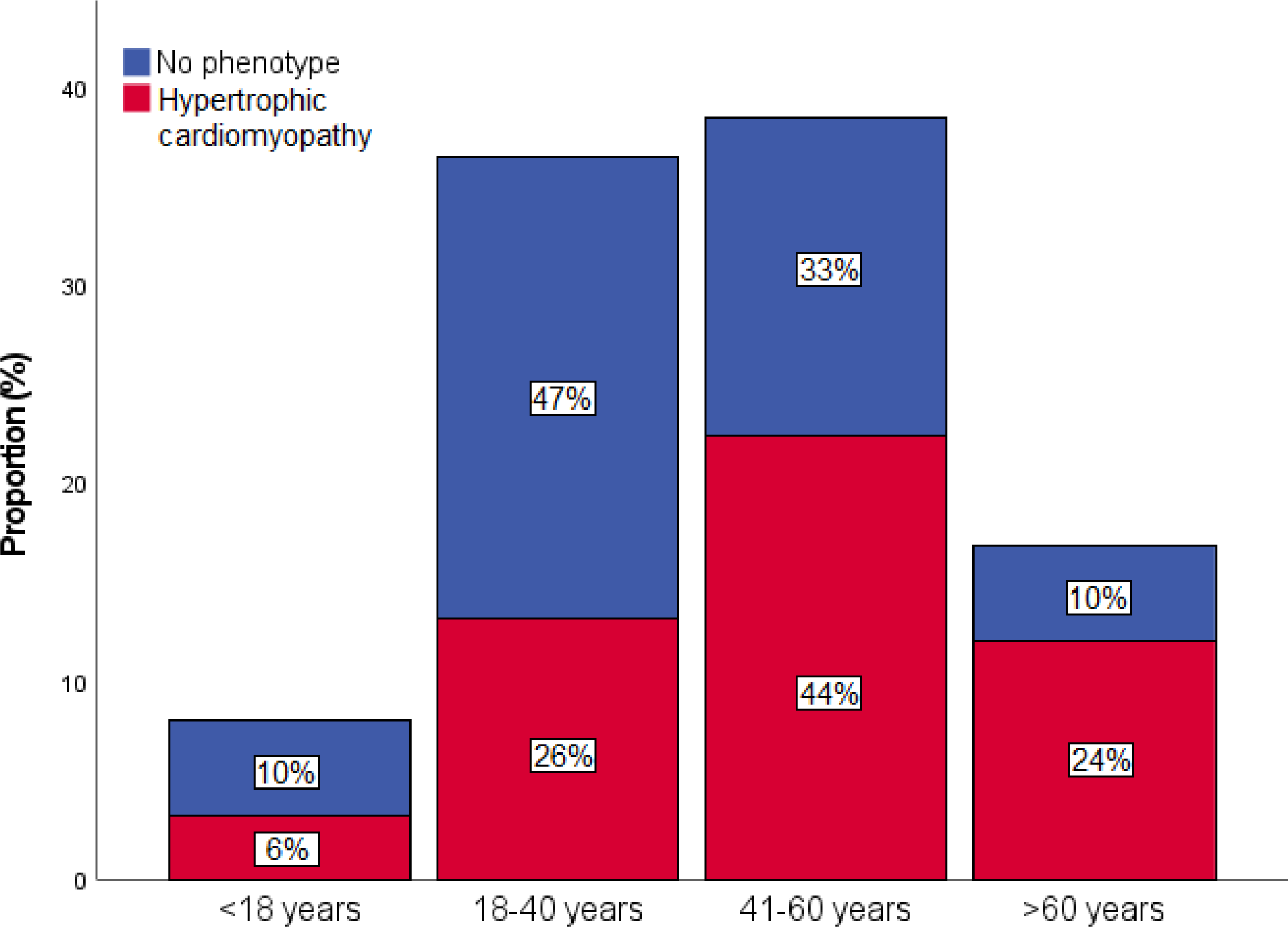
Distribution of genotype-positive individuals without a phenotype and the proportion of genotype-positive individuals diagnosed with hypertrophic cardiomyopathy during their initial cardiologic screening following pre-symptomatic testing, categorized by age group. Percentages indicate the relative distribution within its group.

**Table 1.** Baseline characteristics of genotype positive individuals referred for cardiac screening. Individuals are divided between those who were diagnosed with HCM at first assessment and those without HCM (G+/P-). <u>Abbreviations</u>: ICD: implantable cardioverter-defibrillator; LV: left ventricular; LVOT: left ventricular outflow tract.

| Baseline characteristics upon<br>cardiologic evaluation | Phenotype<br>(HCM; n=128) | No phenotype<br>(G+/P-; n=145) | P value |
| --- | --- | --- | --- |
| Age (years) | 48 (±16) | 38 (±18) | **<.001** |
| Male gender (%) | 73 (57) | 50 (34) | **<.001** |
| Body mass index (kg/m<sup>2</sup>) | 26.3 (±5.4) | 25.1 (±5.4) | 0.069 |
| Hypertension (%) | 29 (23) | 13 (9) | **0.002** |
| **Genotype (%)** |  |  | 0.162 |
| *MYBPC3* | 106 (84) | 108 (76) |  |
| *MYH7* | 12 (9) | 11 (8) |  |
| *MYL2* | 2 (2) | 3 (2) |  |
| *MYL3* | 1 (1) | 1 (1) |  |
| *TNNI3* | 1 (1) | 2 (1) |  |
| *TNNT2* | 0 | 5 (3) |  |
| Other | 6 (3) | 15 (9) |  |
| **HCM-specific characteristics** |  |  |  |
| Left atrial diameter (mm) | 40.0 (±6.2) | 35.5 (±4.6) | **<.001** |
| Intraventricular septum (mm) | 14.7 (±2.6) | 9.3 (±1.7) | **<.001** |
| LV ejection fraction <50% (%) | 0 | 0 | - |
| LVOT gradient, rest (mmHg) | 9.2 (±11.3) | 5.4 (±2.0) | **0.009** |
| LVOT gradient, Valsalva (mmHg) | 17.3 (±20.4) | 7.3 (±4.5) | **0.010** |
| LVOT ≥30 mmHg (%) | 5 (4) | 0 | **0.016** |
| **Outcomes** |  |  |  |
| Follow-up time (years) | 11 (6-16) | 9 (4-14) | **0.014** |
| All-cause death (%) | 14 (11) | 5 (3) | **0.015** |
| Sudden cardiac death (%) | 2 (14) | 0 | 0.372 |
| Time to death (years) | 9 (±3) | 9 (±6) | 0.894 |
| Atrial fibrillation (%) | 15 (12) | 1 (1) | **<.001** |
| ICD (%) | 17 (13) | - | - |
| Septal reduction therapy (%) | 4 (3) | - | - |

### Follow-up of HCM patients

During a follow-up period of 9±4 years, 35 (27%) HCM patients experienced an adverse cardiac event. Among HCM patients, 14 (11%) died: two (14%) from sudden cardiac death, seven (50%) from non-cardiac causes, and five (36%) from unknown (ie, not retrievable) causes. Atrial fibrillation developed in 15 (12%) HCM patients. Four (3%) underwent surgical myectomy. An ICD was implanted for primary prevention in 15 patients (88%), and for secondary prevention in two patients (12%); one following an out-of-hospital cardiac arrest due to ventricular fibrillation, and another after recurrent ventricular fibrillation more than 48 hours post-myocardial infarction. During follow-up, four (24%) of the ICD recipients received appropriate therapy, including two with a primary prevention ICD. Higher age at HCM diagnosis was associated with lower likelihood of surgical myectomy (OR 0.90, 95%CI 0.84-0.97, p=0.006), ICD implantation (OR 0.96, 95%CI 0.93-0.99, p=0.006) and appropriate ICD therapy (OR 0.92, 95%CI 0.86-0.99, p=0.016), per year of diagnosis. Age of diagnosis was not associated with onset of atrial fibrillation (OR 1.03, 95%CI 0.99-1.07, p=0.226). Adverse cardiac outcomes were not associated with specific genes. Patients with an *MYH7* variant (4 out of 12) were not more likely to experience an adverse cardiac outcome than those with an *MYBPC3* (29 out of 105) or other P/LP gene (2 out of 9) variants (33% vs 27% vs 22%, respectively, p=0.710).

### Follow-up of G+/P- individuals

Follow-up data of at least one year were available for 118 out of 145 (81%) G+/P- individuals (Table 2). This cohort had a mean age of 39 years at baseline and 47 years at last follow-up, with a median follow-up time of 8 (Q1-Q3 4-14, range 1-18) years. The majority of this cohort was female (80, 68%). Most individuals (90, 76%) had a P/LP *MYBPC3* variant, followed by 14 individuals (12%) with a P/LP *MYH7* variant and 14 individuals (12%) with a variant in another HCM-associated sarcomeric gene. Two individuals with a P/LP *MYBPC3* variant had a second P/LP variant, both in *MYL2*.

**Table 2.** Echocardiographic parameters of genotype-positive phenotype-negative individuals with follow-up (n = 118). <u>Abbreviations</u>: LV: left ventricular; LVOT: left ventricular outflow tract; TAPSE: tricuspid annular plane systolic excursion; TR: tricuspid regurgitation.

| **Echocardiographic measurements** | **Baseline** | **Last follow-up** | **P value** | **Average paired difference** |
| --- | --- | --- | --- | --- |
| Age (years) | 38.5 (±13.3) | 47.1 (±13.5) | - | - |
| Body mass index (kg/m<sup>2</sup>) | 25.3 (±5.1) | 26.0 (±5.3) | 0.073 | +1.23 (±4.6) |
| Body surface area (m<sup>2</sup>) | 1.88 (±0.23) | 1.91 (±0.23) | 0.086 | +0.03 (±0.46) |
| Left atrial diameter (mm) | 35.0 (±4.5) | 35.7 (±5.7) | 0.088 | +0.68 (±4.3) |
| Left atrial volume index (mL/m<sup>2</sup>) | 18.2 (±9.1) | 22.1 (±8.4) | 0.084 | +2.5 (±7.9) |
| Intraventricular septum (mm) | 9.1 (±1.7) | 9.4 (±1.9) | **0.044** | +0.3 (±1.7) |
| LV posterior wall (mm) | 8.3 (±1.5) | 7.6 (±1.3) | **<.001** | -0.7 (±1.6) |
| Max LV wall thickness (mm) | 9.6 (±1.6) | 9.8 (±1.8) | 0.051 | +0.3 (±1.4) |
| LV end-diastolic diameter (mm) | 46.8 (±4.3) | 46.1 (±4.9) | 0.080 | -0.6 (±3.8) |
| LV end-systolic diameter (mm) | 29.6 (±3.8) | 29.3 (±3.8) | 0.651 | -0.2 (±4.0) |
| TAPSE (mm) | 25.4 (±6.1) | 25.2 (±4.8) | 0.929 | - 0.1 (±5.8) |
| E (m/s) | 0.78 (±0.18) | 0.74 (±0.17) | **0.001** | -0.05 (±0.15) |
| A (m/s) | 0.57 (±0.18) | 0.64 (±0.19) | **<.001** | +0.07 (±0.13) |
| E/A (ratio) | 1.43 (1.05-1.80) | 1.17 (0.90-1.54) | **<.001** | -0.26 (0.48-0.03) |
| Deceleration time (ms) | 190 (±47) | 203 (±51) | **0.031** | +13 (±64) |
| Septal e' (cm/s) | 10.1 (±2.5) | 9.4 (±2.5) | **0.025** | -0.5 (±2.3) |
| Septal E/e' (ratio) | 8.0 (±1.9) | 8.7 (±6.0) | 0.279 | +0.7 (±6.5) |
| TR velocity (m/s) | 1.2 (±1.1) | 1.9 (±2.5) | 0.064 | +0.9 (±3.0) |
| LV ejection fraction (%) | 62.9 (±3.4) | 61.8 (±4.2) | 0.105 | -0.9 (±4.3) |
| Maximal LVOT gradient (mmHg) | 5.1 (±2.4) | 5.7 (±2.5) | **0.021** | +0.7 (±2.9) |

HCM developed in seven (6%) G+/P- individuals after a median time of six years (Q1-Q3 5-16 years), without significant ECG changes occurring in these individuals. These individuals (five, 71% female) were 48±13 years at baseline and 58±11 years upon diagnosis (range 39-77). Mean MWT increased from 10.2±1.2 mm to 13.3±0.5mm at follow-up. Individuals who developed HCM were older at baseline than those who did not (48 vs 38 years, *p*=0.036), were more likely to have a history of hypertension (43% vs 6%, *p*<0.001), had a higher BMI when diagnosed (30.1 vs 25.5 kg/m^2^, *p*=0.033), and on baseline echocardiography had a higher MWT (10.9 vs 9.5 mm, *p*=0.033) and lower septal e’ velocities (8.0 vs 10.2 cm/s, *p*=0.033). One of the individuals who developed HCM died at the age of 78 from a non-cardiac cause, with no other adverse events observed in the remaining six individuals.

Three G+/P- individuals, who did not develop HCM, showed significant ECG changes at last follow-up (one first degree AV block (PR 212 ms), one atrial fibrillation and a right bundle branch block, and one ST-segment depressions with T-wave inversions in leads V3-V6). In this last individual cardiovascular magnetic resonance ruled out (apical) HCM. As shown in Table 2, follow-up measurements revealed significant but clinically negligible mean increases in the intraventricular septum (+0.3 mm, *p*=0.044). In the overall cohort, 66 individuals (56%) exhibited no change (0 mm) in maximal wall thickness measurements, while 24 individuals (20%) had a median increase of 1 mm, 19 individuals (16%) experienced a median increase of 2 mm, and 9 individuals (8%) demonstrated a median increase of 3–5 mm. Among those with a follow-up period of at least five years (n=78), 38 (49%) had a stable (0 mm change) measurement, 17 (22%) individuals had a 1 mm increase, 16 individuals (19%) had an increase of 2 mm, of which three individuals (19%) developed HCM. Additionally, four individuals (5%) exhibited a 3 mm increase, with one individual (25%) developing HCM. Two individuals (3%) had a 4 mm increase, and one individual exhibited a 5 mm increase, with these last three cases all progressing to HCM. The last individual who developed HCM, with a follow-up time of 2 years, did so based on a 1 mm increase (12 to 13 mm). Furthermore, slight decreases were observed in the LV posterior wall (-0.7 mm, *p*<0.001) thickness, the E/A ratio (-0.26, *p*<0.001) and the septal e’ velocity (-0.5 cm/s, *p*=0.025). No individuals developed significant valvulopathy. No individuals developed LVOT obstruction. No individuals had diastolic dysfunction at baseline. Only one individual had developed diastolic dysfunction grade I at the last follow-up (follow-up time of 17 years, age at baseline 49 years). During follow-up, one individual died from a non-cardiac cause. Two G+/P- individuals, both with a pathogenic *MYBPC3* variant, developed paroxysmal atrial fibrillation despite the absence of an HCM phenotype, with one case having occurred prior to baseline assessment. No other adverse cardiac outcomes were observed.

## Discussion

In this study, we evaluated the prevalence of HCM in individuals referred for cardiac screening after PST identified a P/LP variant in a sarcomeric gene. We conducted a longitudinal assessment of echocardiographic changes in those without an initial phenotype. HCM was diagnosed at the first cardiac evaluation in almost half of the G+ individuals, with the age at diagnosis averaging 48 years (range 10-82 years). Individuals diagnosed with HCM upon initial screening were, on average, 10 years older and more likely to be male than G+/P- individuals, consistent with previous findings of age-related disease penetrance and male-dominant prevalence in HCM. [4, 23-25] During a median follow-up period of eight years, only 6% of these individuals developed mild HCM. The median follow-up time to HCM diagnosis was six years. We observed minimal longitudinal echocardiographic changes in individuals who did not develop a phenotype, and the majority had stable a MWT measurement during follow-up.

The high diagnostic yield from initial cardiologic assessment is comparable with previous research, showing a diagnostic rate of approximately 40%. [10, 18] Given that these patients are asymptomatic, they would not have been diagnosed with HCM without PST. Although infrequent, given the occurrence of important cardiac outcomes in this cohort, it highlights the benefit of early identification through PST. These benefits should be weighed against overall low psychological burdens associated with G+ status in those who do not develop adverse outcomes. [26] In those diagnosed with HCM, risk stratification should be performed according to the established guidelines. [7, 8, 27] After an average follow-up of 9 years, a quarter of this HCM cohort experienced an adverse cardiac event, most commonly the implantation of a primary prevention ICD or the development of atrial fibrillation. Younger patients in this cohort were more likely to undergo septal reduction therapy, ICD implantation, and receive appropriate ICD therapy. This aligns with earlier research showing that an earlier age of diagnosis is associated with more severe phenotypes and worse outcomes. [28] Previous research has shown that HCM patients with *MYH7* variants tend to have more severe disease progression and worse outcomes. [29, 30] In this cohort, genotype was not associated with a relative increased risk of adverse cardiac outcomes, although this may have been the consequence of small sample sizes of non-*MYBPC3* variants. The youngest HCM patient identified in our study was 10 years old, and all patients under the age of 16 were carriers of a *MYBPC3* variant. Current European guidelines on family screening advise offering genetic and clinical screening to first-degree relatives starting at the age of 10, whereas American guidelines recommend pediatric screening at the time of HCM diagnosis in family members, and no later than puberty. [7, 8] Earlier testing may be warranted in the presence of cardiac symptoms, a family history of early-onset HCM or sudden cardiac death, or participation in competitive sports. [31, 32] Looking ahead, novel treatments for symptomatic HCM, such as cardiac myosin inhibitors, may promote beneficial cardiac remodeling and potentially prevent adverse changes. Early use of these therapies, following the identification of HCM, could lead to improved outcomes, which would further emphasize the importance of early detection through PST. [33-35] Therefore, future research should investigate whether early treatment with cardiac myosin inhibitors in asymptomatic HCM patients may improve outcomes, given the increased incidence of adverse events in this population and the higher rate of adverse outcomes in younger patients.

The low incidence of HCM development within this G+/P- cohort is consistent with earlier research, which has reported similarly limited phenotype development in those individuals. [4-6, 36] A recent report by Silajdzija et al. in 2024 reported a comparable 4% diagnostic yield during a seven-year follow-up period. Earlier research has suggested that diastolic functional abnormalities can precede the development of LVH. [37] However, in individuals who did not develop LVH during follow-up, echocardiographic changes were subtle, with minimal changes in wall thickness and the development of most likely age-related mild diastolic dysfunction in only one individual. Additionally, a previous study demonstrated that LV parameters, including diastolic function, in G+/P- individuals are not significantly different compared to those in healthy controls. [38] Overall, the observed changes in LV and diastolic function measurements in our study were negligible and do not indicate accelerated negative remodeling or diastolic dysfunction attributable to pre-phenotypical HCM-related changes when compared to the general population. [39] While ECG changes, particularly the development of Q waves, have been implicated as predictors of phenotypic development, this study found that significant ECG changes were infrequent in the individuals not prone to phenotype development. [14, 18, 36, 40] Adverse cardiac events, including cardiac death, did not occur. The low incidence of events supports a lower follow-up frequency in G+/P- individuals, with an interval of at least 5 years appearing safe and reasonable.

Individuals diagnosed with HCM had a higher prevalence of hypertension. Although no causal relationship has been established between arterial hypertension and HCM, this finding is noteworthy, as previous data have shown an association between arterial hypertension and HCM in genotype-negative patients. [41] Furthermore, those who developed a phenotype during their follow-up had a significantly higher BMI, in line with previous research linking higher BMI to adverse LV remodeling and changes in cardiomyocyte structure and metabolism in HCM patients. [42, 43] Future research should explore whether cardiovascular risk factors, such as arterial hypertension and obesity, independently contribute to the development of the HCM phenotype, potentially as a second-hit phenomenon. If such mechanisms are confirmed, more aggressive management strategies for hypertension and overweight to prevent HCM development in susceptible individuals might be warranted. A recent randomized controlled trial (VANISH) investigated whether valsartan, an angiotensin II receptor blocker, could modify disease progression in subclinical HCM patients, but it did not show an effect on phenotype progression. [44] However, this trial did not include individuals with hypertension.

This retrospective study had limitations. Besides for *MYBPC3* variants, the sample size for other P/LP variants was limited, potentially affecting the robustness of genotype-specific comparisons. Minor differences in measurements could have been caused due to inter-user variability and changes in echocardiography hardware over time. However, all images were reexamined to minimize this variability. Additionally, the varying follow-up times further complicate cohort comparisons. Future prospective research on G+/P- individuals should include longitudinal assessments to evaluate precise phenotypical changes over time and attempt to correlate these changes with secondary factors such as environmental influences and cardiovascular health and risk factors (e.g., arterial hypertension and obesity).

In conclusion, baseline evaluation following PST revealed a high diagnostic yield, identifying HCM in about half of the G+ individuals. In G+/P- individuals, only minimal longitudinal echocardiographic changes and infrequent phenotype development were observed. Therefore, a reduced follow-up frequency (eg, every 5 years) is appropriate for these individuals.

## Data Availability

The data that support the findings of this study are available from the corresponding author, SACS, upon reasonable request.

## Non-standard Abbreviations and Acronyms

ECG: electrocardiogram
G+/P-: genotype-positive, phenotype negative
HCM: Hypertrophic cardiomyopathy
ICD: implantable cardioverter-defibrillator
LVH: left ventricular hypertrophy
LVOT: left ventricular outflow tract
MWT: maximal wall thickness
P/LP: pathogenic/likely pathogenic gene variant
PST: pre-symptomatic testing

## Acknoledgment

This paper is part of the project ‘Double Dose of energy and efforts of the national DOSIS consortium to design and test new diagnostic and treatment strategies for inherited cardiomyopathies (DOUBLE DOSE), which is funded by the Dutch Heart Foundation and the Dutch Foundation (Stichting) Hartedroom (Grant number: 2020B005).

